# Effectiveness and Types of Interventions for Autism Spectrum Disorder: A Systematic Review and Meta-Analysis

**DOI:** 10.64898/2026.04.28.26351945

**Authors:** John K. Muthuka, Ruvimbo Zimunya, Andrina Simengwa, Chrisphine Onyango, Kelly Oluoch, Mary T. Kioko, Diana F. Mbari, Japheth M. Nzioki, Lucy K. Chebungei, Sara Kim, Desire A. Nshimirimana

## Abstract

This systematic review and meta-analysis aimed to estimate the overall effectiveness of ASD interventions and identify sources of heterogeneity using frequentist and Bayesian approaches. A systematic search of PubMed/MEDLINE, Embase, Web of Science, and Scopus was conducted for studies published between January 1, 2004, and April 30, 2025. Mainly, randomized controlled trial studies with extractable intervention outcomes were included. A total of 41 studies (n=3,008) were synthesized using random-effects models (REML), Bayesian hierarchical modeling, meta-regression, and sensitivity analyses following PRISMA guidelines. The pooled random-effects estimate showed a significant positive effect of ASD interventions (effect size = 0.506, 95% CI: 0.392–0.619; z = 8.72, p < .001), corresponding to an estimated success proportion of 62% (95% CI: 59%–65%). Heterogeneity was substantial (Q□(40) = 238.78, p < .001; I² = 82.45%; τ² = 0.069, 95% CI: 0.028–0.137; τ = 0.262), with H² = 5.70 and a wide prediction interval (−0.020 to 1.031), indicating strong between-study variability. Bayesian meta-analysis confirmed a comparable effect (posterior mean = 0.619, 95% CrI: 0.592–0.646), with τ = 0.273 and I² ≈ 82.5%, and MCMC diagnostics showed stable convergence (R-hat ≈ 1.00). Publication bias analyses indicated significant funnel plot asymmetry (Egger-type regression: z = 3.429, p < .001; weighted regression: t = 9.573, p < .001), while rank correlation was non-significant (τ = −0.178, p = .103). Trim-and-fill analysis imputed 10 studies, reducing the pooled effect to 0.374 (95% CI: 0.258–0.491; τ = 0.338), though the effect remained significant (p < .001). Sensitivity analyses excluding influential studies yielded a stable effect (0.505, 95% CI: 0.401–0.609), with persistent heterogeneity (I² = 75.49%; Q□(38) = 190.21, p < .001; τ² = 0.043). Subgroup analyses showed highest effects for digital/technology-based interventions (0.672; 67%; I² = 0%), followed by nutritional (0.635; 64%; I² = 73.81%), behavioral (0.630; 63%; I² = 74.78%), and pharmacological (0.627; 63%; I² = 0%) interventions, while physical/occupational therapies showed lower effects (0.523; 52%; I² = 63.35%) and combined interventions showed borderline effects (0.593; 59%; I² = 19.96%); subgroup differences were significant (Q□5) = 22.63, p < .001). Regional effects were similar and non-significant (Q□(2) = 0.73, p = .694): North America (0.619; I² = 84.21%), Europe (0.626; I² = 62.34%), and Asia (0.659; I² = 0%). Age at intervention onset did not significantly moderate effects (Q□(5) = 0.98, p = .964), although variability was observed across children, adolescents, adults, and toddlers. Meta-regression identified significant moderators including intervention context (Q□= 18.159, p = .020), outcome domain (Q□= 19.588, p = .003), age at start (Q□= 17.795, p = .003), and intervention category (Q□= 31.714, p < .001), while follow-up duration and intervention duration were not significant. Bayesian subgroup analyses confirmed robustness, with strongest evidence for pharmacological (BF → ∞), behavioral (BF ≈ 832.50), and digital interventions (BF ≈ 30.92). In conclusion, ASD interventions demonstrate a moderate and statistically significant overall effect (∼0.50–0.62), with substantial heterogeneity driven primarily by intervention type, context, and participant characteristics, and findings were consistent across frequentist, Bayesian, and sensitivity analyses, supporting robust but context-dependent effectiveness.

## Introduction and Background

Autism Spectrum Disorder (ASD) is a neurodevelopmental condition characterized by persistent differences in social communication and restricted, repetitive patterns of behavior that typically emerge in early childhood ^1–5^. The global burden of ASD has become an increasing public health priority due to its lifelong functional implications and substantial social and economic costs ^6^. While accurate prevalence estimates are essential for informing health policy and guiding early detection strategies, equally critical is the evaluation of intervention effectiveness, given the need to improve developmental outcomes and reduce disability across diverse populations.

Over the past two decades, a wide range of interventions—including behavioral, developmental, educational, and pharmacological approaches—have been implemented worldwide. Evidence from randomized controlled trials and systematic reviews suggests that structured behavioral interventions, such as Applied Behavior Analysis (ABA) and Naturalistic Developmental Behavioral Interventions (NDBI), consistently improve communication, adaptive functioning, and cognitive skills ^7,8^. Broader early childhood programs and comprehensive developmental interventions have also demonstrated benefits, though their effects are more variable and context-dependent ^9^. Despite these advances, substantial heterogeneity in reported outcomes persists, reflecting differences in study design, intervention intensity, participant characteristics, and healthcare infrastructure.

Previous meta-analyses have provided pooled estimates of intervention effectiveness, but many have focused narrowly on specific modalities or regions, leaving gaps in understanding the broader drivers of variability. Given ongoing changes in diagnostic practices, awareness, and intervention delivery from 2004 to 2025, an updated synthesis is needed to quantify both the overall effectiveness of autism interventions and the methodological and contextual moderators contributing to heterogeneity.

The primary objective of this study was therefore to evaluate both pooled and specific effectiveness of various autism interventions across diverse populations; and second, to identify and quantify the study-level drivers of between-study heterogeneity using Bayesian meta-analytic and meta-regression techniques. This dual focus provides not only a precise estimate of intervention impact but also critical insights into the conditions under which interventions are most effective, thereby informing clinical practice, policy development, and future research.

## Methods

### Study design and protocol registration

A systematic review and meta-analysis of ASD interventions studies was carried out. The study adhered to the Preferred Reporting Items for Systematic Reviews and Meta-Analyses guidelines ^10,11^. The current review is part of a larger, prospectively registered study titled *Global Trends in Autistic Disorder Diagnosis and Intervention Outcomes-A Systematic Review, Meta-analysis, and Meta-Regression* (PROSPERO: CRD420251069271), with a particular emphasis on evaluating the effect of the ASD interventions cumulatively and also, specifically as per the different regions of the globe, spanning 2004 to 2025

### Search strategy and study selection

Our systematic review and meta-analysis included studies published between January 1, 2004, and April 30, 2025, that evaluated the effectiveness and types of interventions for autism spectrum disorder (ASD). Eligible studies were required to assess at least one intervention targeting individuals with ASD and report measurable outcomes related to effectiveness. We included randomized controlled trials, quasi-experimental studies, and observational designs, provided that extractable outcome data were available. Interventions delivered in clinical, educational, community, or home-based settings were considered. We excluded studies that addressed non-ASD populations, used duplicate datasets, or were case reports, case series, reviews, meta-analyses, editorials, or commentaries. Studies without full-text availability or without clearly defined intervention methods or outcome measures were also excluded. Additionally, genetic, neurobiological, or laboratory-based studies that did not evaluate intervention effectiveness were not considered. A comprehensive electronic search was conducted in PubMed/MEDLINE, Embase, Web of Science, and Scopus for studies published between January 1, 2004, and April 30, 2025. Grey literature sources were also searched. The search strategy combined controlled vocabulary (Medical Subject Headings [MeSH]) and free-text keywords such as “autism spectrum disorder,” “intervention,” “therapy,” and “treatment” using Boolean operators (AND/OR). No language or geographic restrictions were applied.

### Eligibility criteria

This systematic review and meta-analysis included RCT studies documenting the type of ASD intervention alongside their effect on it (ASD) and were conducted in any part across the globe. Studies were excluded if they were experimental, case series, case reports, letters, opinions, narrative reviews, or other studies that did not contain primary data, if they were duplicates, or if they did not have full text. Studies lacking clear diagnostic criteria or denominators were excluded from quantitative synthesis.

### Data extraction

In case of duplicate publications, the article that contained the most information was included in the review and all others were excluded as duplicates. The data extracted from the studies included: publication year, country, study design, study period, ASD screening tool used, intervention type and category, intensity of the intervention, mean age of participants, sample size, outcome domain, follow-up duration and study context (e.g clinic based). For each study, two reviewers independently extracted the data into a standardized data extraction sheet using Microsoft Office Excel. Disparities in data extracted were resolved via discussion between the reviewers.

### Assessment of the quality and risk of bias of included studies

The risk of bias of the included studies was assessed using a domain-based approach appropriate for randomized controlled trials, and the results were visualized using the ROBVIS ^12^. Key domains evaluated included the randomization process, deviations from intended interventions, missing outcome data, measurement of outcomes, and selection of reported results. Each study was judged as having low risk, some concerns, or high risk of bias across these domains. All included studies were independently assessed by two reviewers, with disagreements resolved through discussion or consultation with a third reviewer.

### Synthesis of findings and statistical analysis

We narratively described non-meta analyzed data, which included author names, year of publication, continent, study design, and sample size. Unadjusted effect estimates of ASD interventions and their 95% confidence intervals were calculated for each included study, and they were then pooled, when possible. Meta-analyses were performed using JASP (Version 0.95.4.0)^13–15^ with random-effects models estimated via restricted maximum likelihood (REML). To stabilize variance, ASD intervention’s estimates were logit-transformed and subsequently back-transformed for interpretability. Between-study heterogeneity was assessed using Cochran’s Q statistic, I², and τ². Influence diagnostics and Baujat plots were used to identify studies exerting disproportionate effects on pooled estimates. Funnel plots were generated to evaluate potential small-study effects, with asymmetry interpreted cautiously. Additionally, residual funnel plots adjusted for study-level moderators were examined.

Meta-regression analyses were conducted to examine pre-specified moderators, including intervention context, follow-up duration, outcome domain, age at intervention initiation, intervention duration, and intervention category. For each moderator, initial omnibus tests were performed, followed by examination of individual coefficients to identify specific subgroup differences. Multi-collinearity was assessed using variance inflation factors (VIF)^16^, which indicated substantial overlap among predictors and informed model interpretation and refinement. Model stability was further evaluated through sensitivity analyses and influence diagnostics. These included leave-one-out analyses, exclusion of influential studies, and comparison of model estimates after removing studies contributing to sparse or unstable category estimates, particularly within multi-level categorical moderators. These procedures were used to assess the robustness of both the pooled effect size and the observed moderator effects. In addition, a Bayesian random-effects meta-analysis^17,18^ was performed using weakly informative priors, with a Normal (0,1) prior specified for the pooled logit effect size and a Half-Cauchy (0,1) prior for between-study heterogeneity (τ). Markov Chain Monte Carlo (MCMC) sampling^19–23^ was conducted with appropriate burn-in and iteration settings. Convergence was assessed using R-hat statistics and effective sample sizes. Posterior distributions were examined to evaluate the precision of effect estimates and to confirm that estimates were well-separated from the null, indicating robust evidence of intervention effects.

## Results

### Included Articles and Quality Assessment (Systematic Review)

The database search identified 1,067 records. After removing 537 articles without associated data as deserved, 530 records remained for title and abstract screening. Of these, 281 were excluded, and 249 reports were sought for retrieval. A total of 150 reports could not be retrieved, leaving 99 full-text articles assessed for eligibility. After full-text evaluation, 57 articles were excluded for predefined reasons. Ultimately, 42 studies ^24,25,34–43,26,44–53,27,54–63,28,64,29–33^.met the inclusion criteria and were included in the quantitative synthesis *(Figure 1)*.

**Figure 1:**
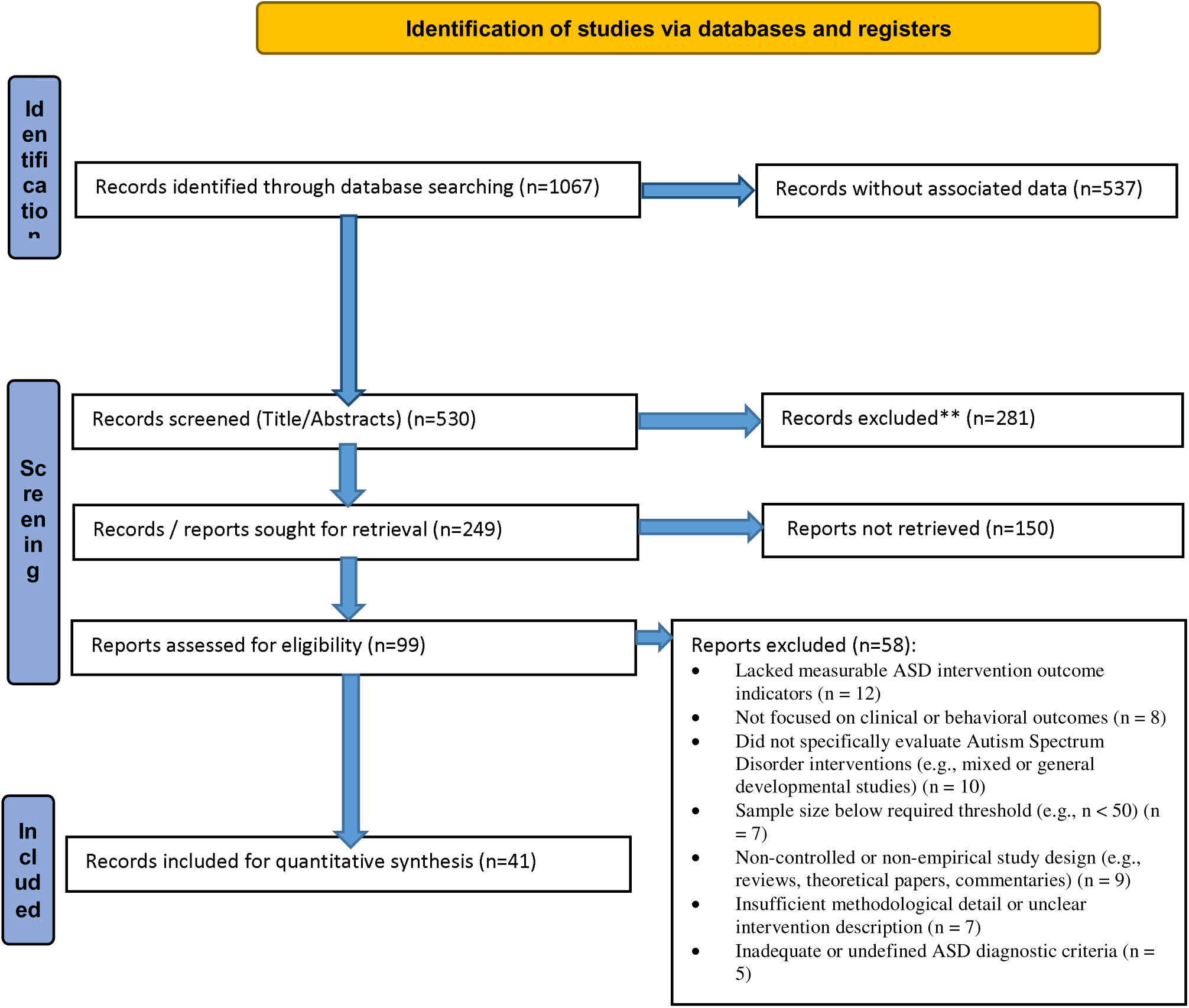
Identification and selection of studies/records in the databases PRISMA: Preferred Reporting Items for Systematic Review and Meta-Analysis

**Figure 2.**
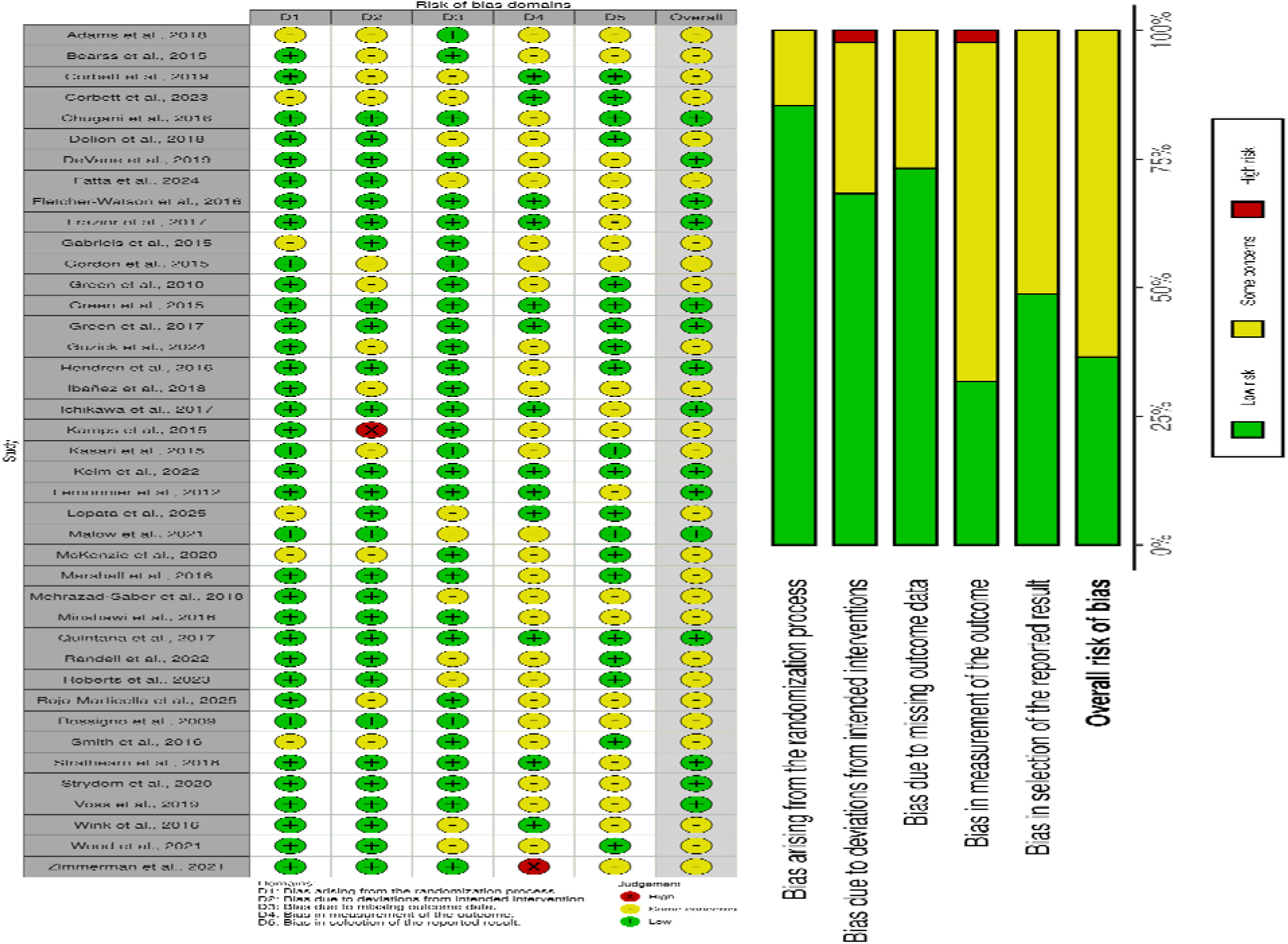
Risk of bias assessment of included studies using the Cochrane Risk of Bias 2 (RoB 2) tool, and visualized using the Robvis tool.

### Features of the Included Studies

The 41 included studies ^24,25,34–43,26,44–53,27,54–63,28,64,29–33^ (n = 3,008) evaluated a wide range of ASD interventions, mainly behavioral and psychosocial approaches^10,25,53,56,57,60,61,63,65,66,32–35,38,40,44,49^, with additional pharmacological interventions^26,28,37,39,43,47,52,55,58,64^, nutritional and biomedical approaches^24,42,50,51,67^, physical and occupational therapies^31,48,59^, digital interventions^29,30,54^, and combined or multimodal approaches^46,62^. Most were randomized controlled trials conducted in North America and Europe, primarily involving children. Interventions were generally short-term (<3 months) and delivered across home, clinic, school, and community settings. Outcomes included ASD symptoms, social communication, behavior, sleep, and adaptive functioning. Follow-up was mostly limited to post-treatment, and overall effects were moderate, with stronger findings observed for behavioral interventions *(Table 1)*.

**Table 1:**
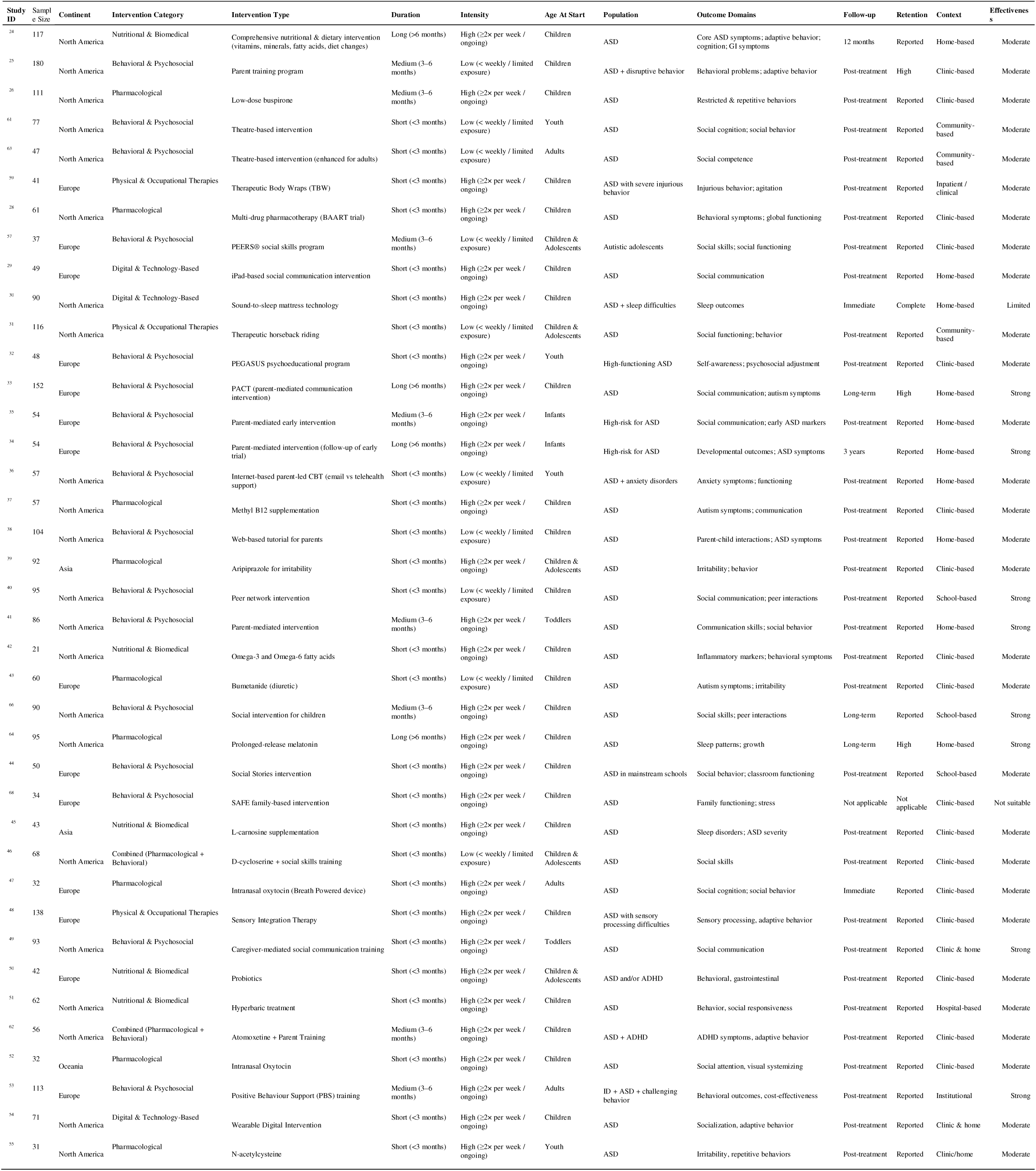
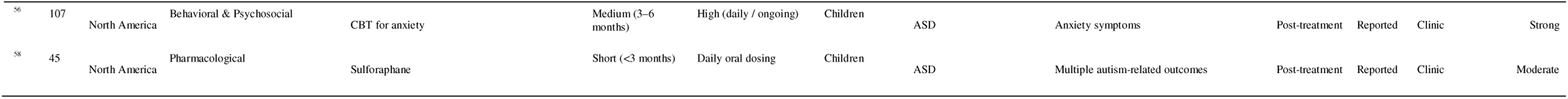
Characteristics of Included Studies on Interventions for Autism Spectrum Disorder.

### Risk of bias assessment

The overall risk of bias across the 41 included studies was moderate, with some variability across domains. Fifteen studies (37%) were rated as having low risk of bias ^26,28,53–55,64,29,34,35,37,39,42,43,47^, reflecting strong methodological rigor, including appropriate randomization, minimal missing data, and reliable outcome measurement. The remaining 26 studies (63%) were rated as having some concerns^24,25,48–52,57–59,61,62,31,63,65–69,32,33,38,40,41,44,46^, Primarily due to deviations form intended interventions, limitations in outcome measurement, and potential selective reporting. Across domains, the randomization process was generally well conducted, and missing outcome data were largely well managed. In contrast, deviations from intended interventions and outcome measurement were the main sources of bias. Overall, the evidence base indicates a predominantly moderate risk of bias, with generally consistent findings suggesting beneficial intervention effects, although the certainty of evidence remains moderate *(Figure2)*.

### Pooled Effect and Heterogeneity of Interventions

A random-effects meta-analysis was conducted to estimate the overall effect of ASD interventions across diverse populations (n = 41) ^24,25,35–44,26,45–54,28,55–64,29,66,30–34^. The overall pooled effect was statistically significant, z = 8.72, p < .001. The pooled effect size was 0.506, 95% CI [0.392, 0.619], corresponding to an estimated proportion of 62% (95% CI [59%, 65%]) when back-transformed to the proportion scale. These results indicate that, on average, a substantial proportion of individuals benefit from the interventions across the populations studied. Heterogeneity among studies was substantial (Q (40) = 238.78, p < .001), with a large between-study variance, τ² = 0.069, 95% CI [0.028, 0.137]; τ = 0.262, 95% CI [0.167, 0.371]. This indicates that the true effect sizes varied considerably across studies, likely due to differences in study populations, intervention types, or implementation contexts. Consistent with this, heterogeneity was high (I² = 82.45%), indicating that most of the observed variability reflects real differences rather than sampling error. The H² statistic further confirmed substantial dispersion (H² = 5.70, 95% CI [2.92, 10.42]), suggesting strong inconsistency across study-level effects. The 95% prediction interval ranged from −0.020 to 1.031 on the effect size scale, indicating that in some future contexts the intervention effect may be minimal or absent, while in others it may be very large. This wide prediction interval highlights the variability and contextual dependence of intervention effectiveness (*Table 2; Figure 3)*.

**Figure 3.**
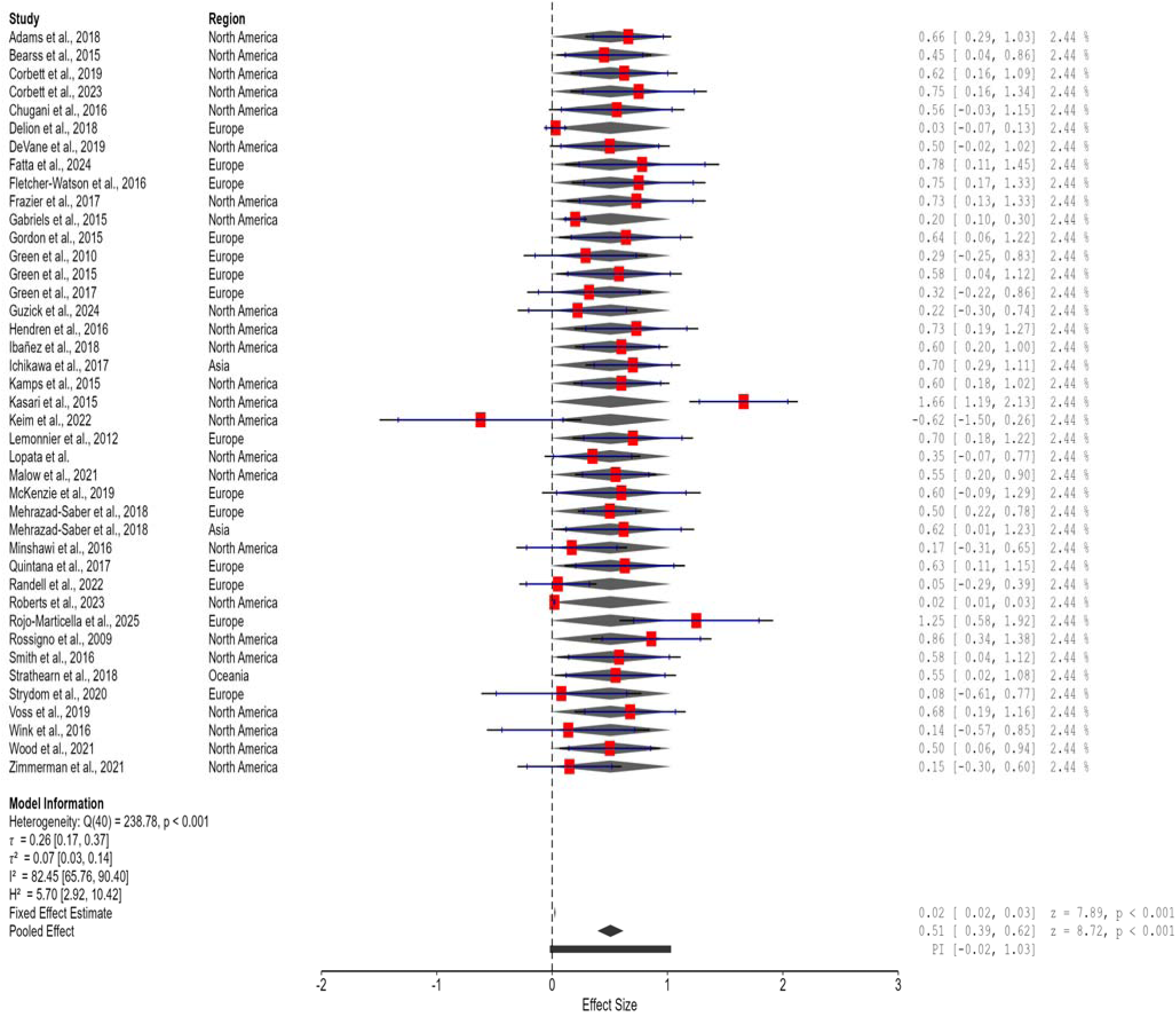
Forest plot of pooled effect of ASD interventions. Forest plot showing a statistically significant pooled effect of ASD interventions across studies (n = 41), based on a random-effects meta-analysis (effect size = 0.506, 95% CI [0.392, 0.619], p < .001). Substantial heterogeneity was observed (I² = 82.45%), indicating variability in effects across studies.

**Table 2.**
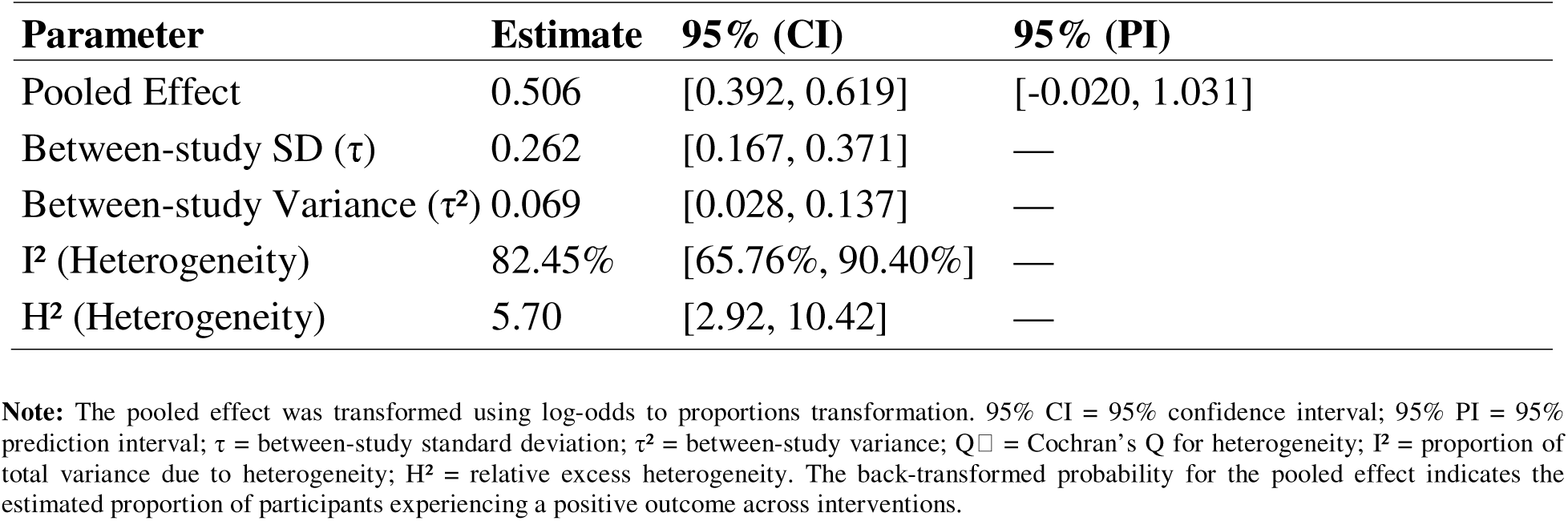
Meta-Analytic Summary of ASD Interventions.

### Bayesian Pooled Meta-Analysis Results and Diagnostics

The Bayesian meta-analysis shows that autism interventions have a moderate-to-large positive effect, with a pooled mean of 0.619 (95% CI: 0.592–0.646) and a similar median, indicating a symmetric distribution. There is substantial variability between studies (I² ≈ 82.5%; τ = 0.273), but the prediction interval (0.481–0.742) suggests future results will still be positive. The posterior is clearly above zero, and the MCMC diagnostics (well-mixed chains, low autocorrelation, stable distributions) indicate reliable estimates. Overall, the findings support that autism interventions are effective despite differences across studies *(Table 3; Figure 4)*.

**Figure 4:**
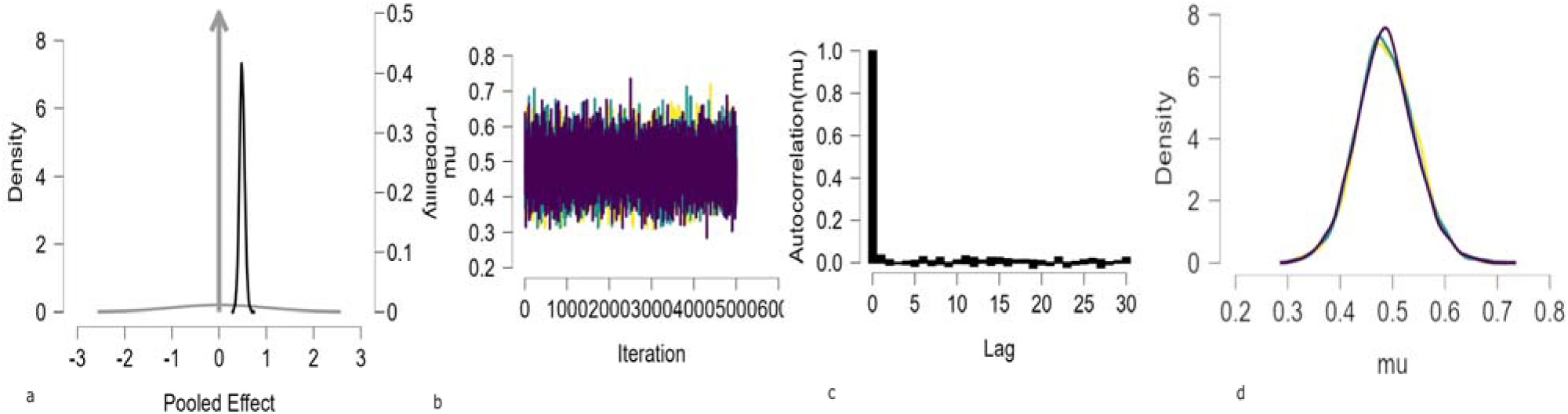
Markov Chain Monte Carlo (MCMC) diagnostic plots for the model parameters: (a) posterior density of the pooled effect (μ), (b) trace plots across multiple chains, (c) autocorrelation function, and (d) posterior density of μ. The pooled effect is positive and precise (μ ≈ 0.62, 95% CI [0.593, 0.647]), although the prediction interval suggests variation across studies. The trace plots show well-mixed, stable chains, indicating good convergence. Autocorrelation decreases rapidly, showing efficient and mostly independent sampling. The posterior distribution is smooth and consistent across chains, while substantial heterogeneity (I² ≈ 82%) indicates notable differences between studies.

**Table 3.**
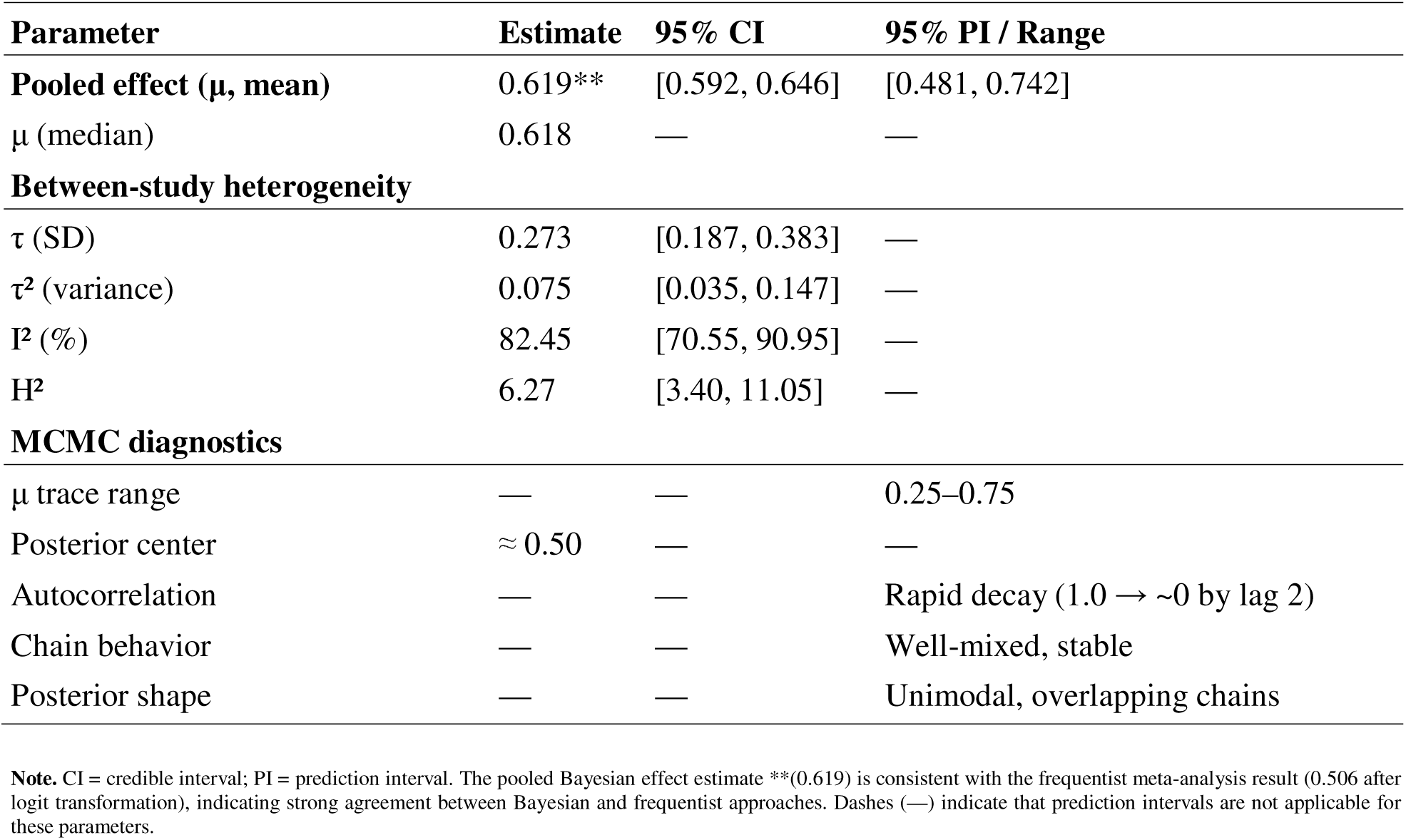
Bayesian Meta-Analytic Estimates for Autism Interventions.

| Parameter | Estimate | 95% CI | 95% PI / Range |
| --- | --- | --- | --- |
| **Pooled effect ($\mu$, mean)** | 0.619** | [0.592, 0.646] | [0.481, 0.742] |
| $\mu$ (median) | 0.618 | — | — |
| **Between-study heterogeneity** |  |  |  |
| $\tau$ (SD) | 0.273 | [0.187, 0.383] | — |
| $\tau^2$ (variance) | 0.075 | [0.035, 0.147] | — |
| $I^2$ (%) | 82.45 | [70.55, 90.95] | — |
| $H^2$ | 6.27 | [3.40, 11.05] | — |
| **MCMC diagnostics** |  |  |  |
| $\mu$ trace range | — | — | 0.25–0.75 |
| Posterior center | $\approx 0.50$ | — | — |
| Autocorrelation | — | — | Rapid decay (1.0 → $\sim 0$ by lag 2) |
| Chain behavior | — | — | Well-mixed, stable |
| Posterior shape | — | — | Unimodal, overlapping chains |

**Note.** CI = credible interval; PI = prediction interval. The pooled Bayesian effect estimate \*\* (0.619) is consistent with the frequentist meta-analysis result (0.506 after logit transformation), indicating strong agreement between Bayesian and frequentist approaches. Dashes (—) indicate that prediction intervals are not applicable for these parameters.

### Assessment of Small-Study Effects and Publication Bias

Funnel plot asymmetry was assessed using three tests, with both the meta-regression (z = 3.429, p < .001) and weighted regression (t = 9.573, df = 39, p < .001) showing significant asymmetry, suggesting possible small-study effects or publication bias, while the rank correlation test was not significant (τ = −0.178, p = .103). Overall, this indicates funnel plot asymmetry, with smaller studies tending to report larger effects. The trim-and-fill analysis further supported potential publication bias by imputing 10 missing studies, which reduced the pooled effect to μ = 0.374 (95% CI [0.258, 0.491], p < .001), although it remained significant. Heterogeneity also remained substantial (τ = 0.338), indicating that while the corrected effect is smaller, a positive effect persists after adjustment (*Table 4; Figure 5*).

**Figure 5:**
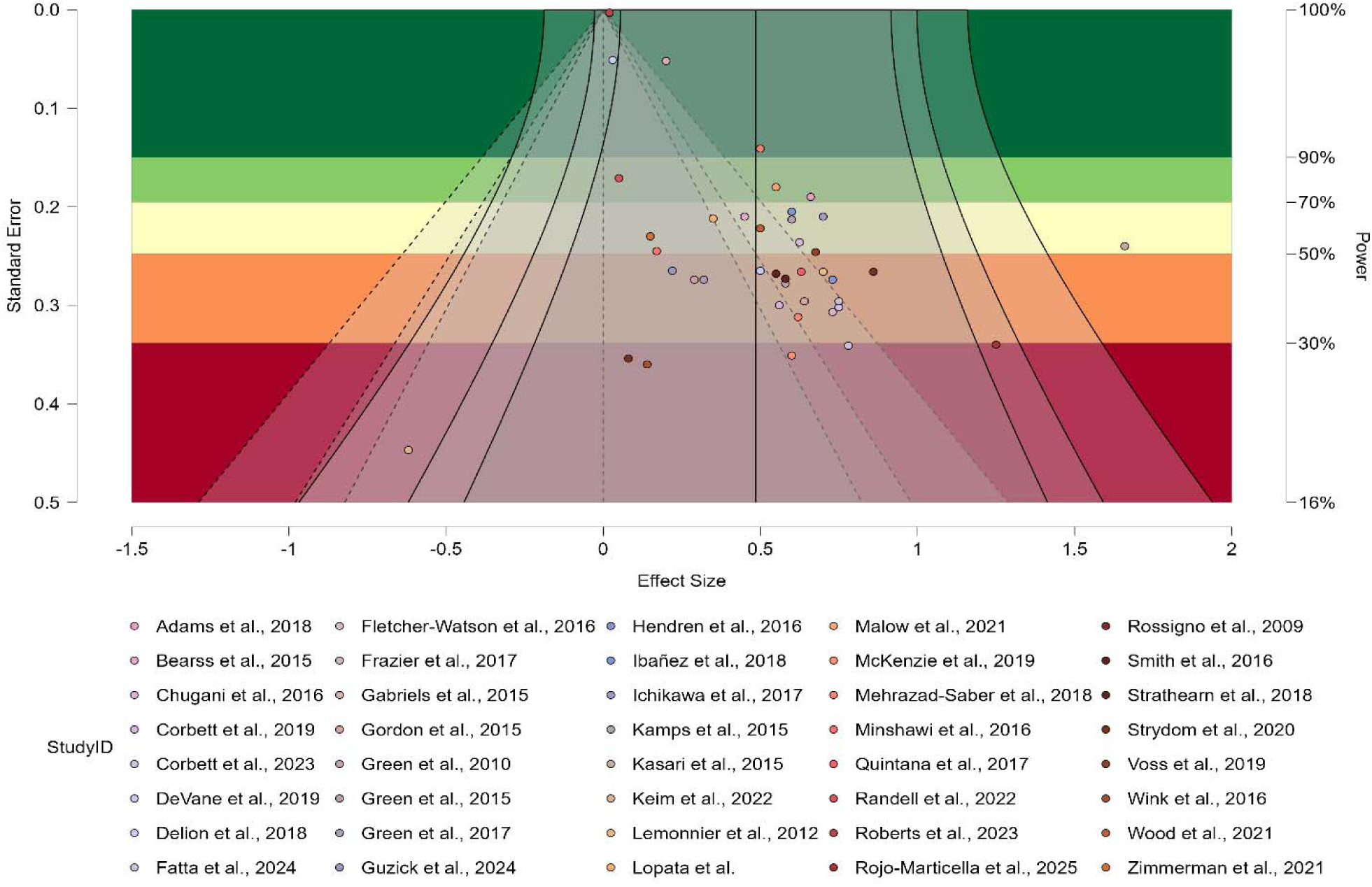
Funnel plot of included studies assessing pooled ASD interventions.

**Table 4.** Funnel Plot Asymmetry Tests.

| Method | Statistic | df | p-value | Estimate ($\mu$) | 95% CI | Missing Studies | Adjusted $\tau$ | 95% CI ($\tau$) |
| --- | --- | --- | --- | --- | --- | --- | --- | --- |
| Meta-regression test | $z = 3.429$ | — | < .001 | 0.129 | [-0.079, 0.337] | — | — | — |
| Weighted regression test | $t = 9.573$ | 39 | < .001 | 0.014 | [0.005, 0.022] | — | — | — |
| Rank correlation test | $\tau = -0.178$ | — | .103 | — | — | — | — | — |
| Trim-and-fill | — | — | < .001 | 0.374 | [0.258, 0.491] | 10 | 0.338 | [0.252, 0.474] |

**Note.** df = degrees of freedom; $\mu$ = limit estimate; CI = confidence interval. The meta-regression and weighted regression tests indicate significant asymmetry, suggesting potential small-study effects, whereas the rank correlation test was not statistically significant. Estimates $\mu$ and $\tau$ are on the logit-transformed prevalence scale. Meta-regression and weighted regression tests assess funnel plot asymmetry; non-significant p-values indicate minimal evidence of publication bias.

### Diagnostic Evaluation of Heterogeneity and Publication Bias

To assess the robustness of the pooled effect and explore potential sources of heterogeneity, two diagnostic plots were generated. The Baujat plot showed that most studies contributed little to overall heterogeneity, but a small subset ^41,42^ exerted disproportionate influence on both variability and the pooled effect estimate. In parallel, the funnel plot indicated that while the majority of studies were symmetrically distributed within the funnel boundaries, a few lay outside ^41,42^, suggesting mild asymmetry. Taken together, these diagnostics suggest that heterogeneity is largely driven by a handful of influential studies, and although some asymmetry is present, the overall pooled effect remains robust, with deviations more consistent with genuine variability than with widespread publication bias *(Figure 6)*.

**Figure 6.**
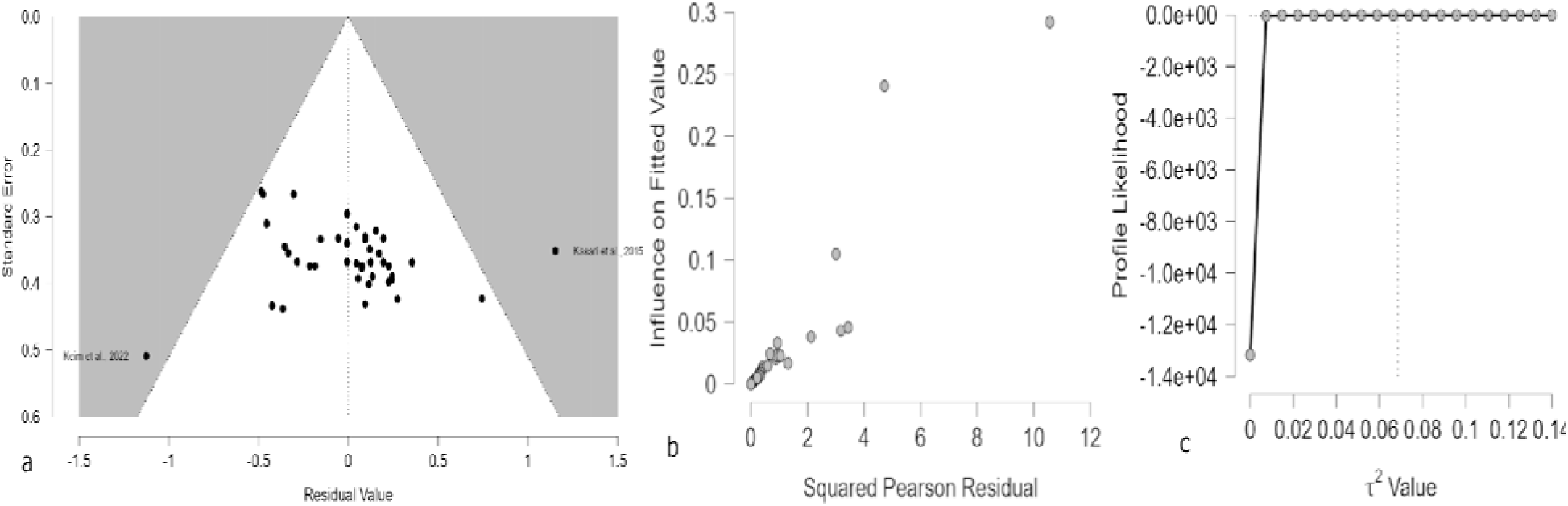
(a) Residual funnel plot illustrating the distribution of standardized residuals against their standard errors to assess small-study effects and potential publication bias; (b) Baujat plot displaying the contribution of individual studies to overall heterogeneity and their influence on the pooled effect size; and (c) profile likelihood plot for τ², depicting the estimation of between-study variance and its associated uncertainty.

**Figure 6a:**
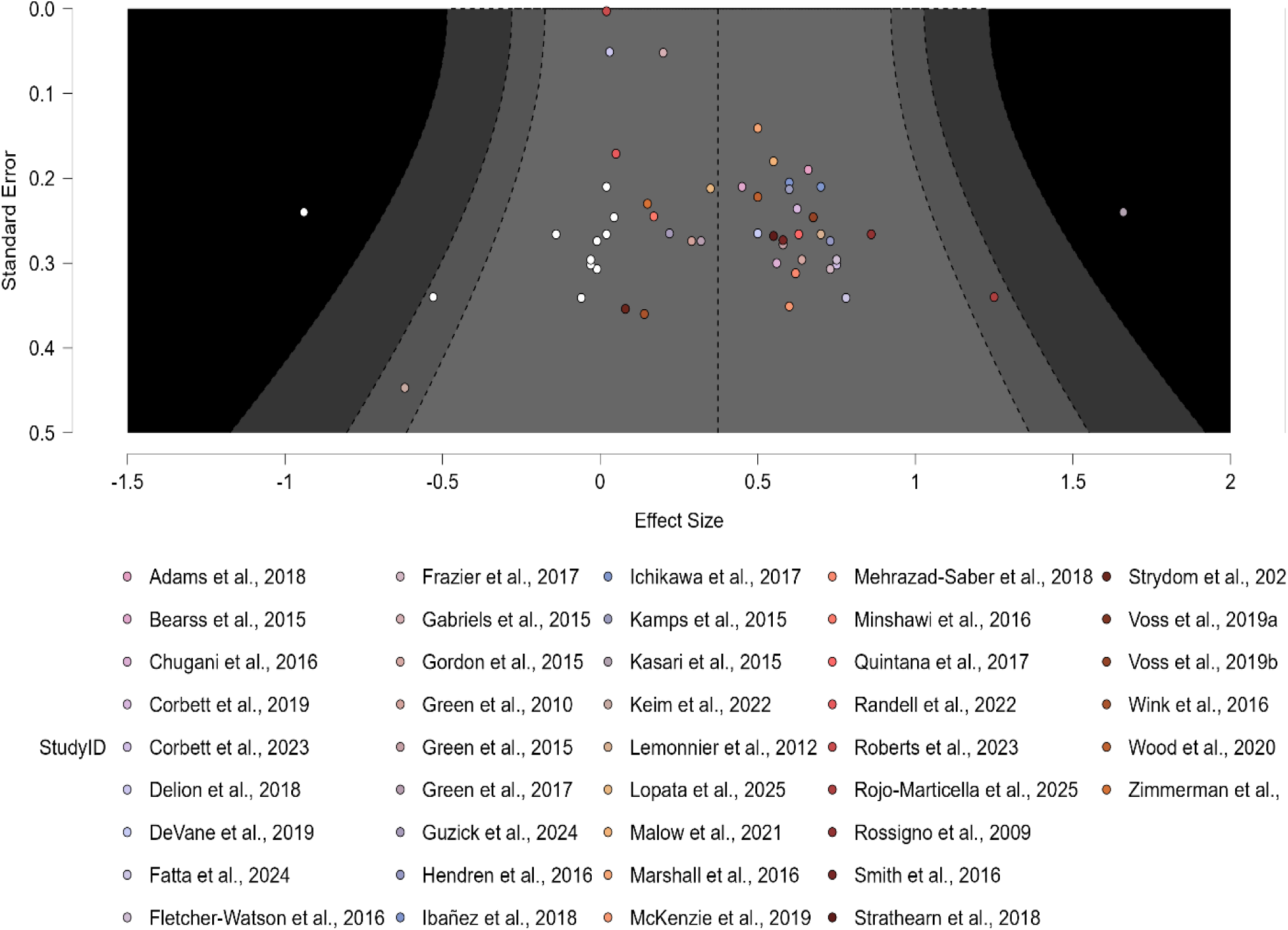
Funnel plot after the Trim and Fill adjustment, illustrating the correction for publication bias and a more symmetrical distribution of studies.

### Assessment of Funnel Plot Asymmetry and the Impact of Trim and Fill Adjustment on Effect Size Estimates

The random□effects meta□analysis yielded a pooled logit effect of 0.506, 95% CI [0.392, 0.619], corresponding to an estimated proportion of 62% (95% CI [59%, 65%]) when back-transformed to the proportion scale. Application of the trim-and-fill method suggested 10 potentially missing studies, producing an adjusted pooled estimate of 0.374 (95% CI: 0.258–0.491, p < .001) for μ, and 0.338 (95% CI: 0.252–0.474, p < .001) for τ. The back-transformed estimate corresponds to an approximate probability of 59%, indicating a comparable magnitude of effect after adjustment. Both the original and adjusted estimates remained statistically significant, suggesting the robustness of the intervention effect even after accounting for potential publication bias. Overall, while the observed effect size may be slightly inflated, the findings indicate that ASD interventions consistently demonstrate meaningful benefits across diverse populations (*Table 5, Figure 6)*.

**Table 5.**
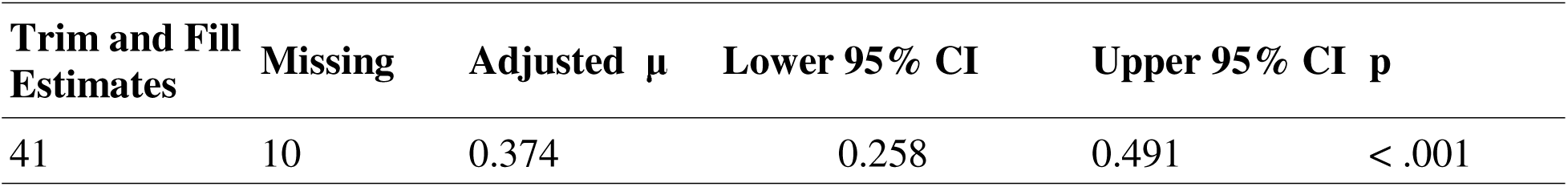
Trim and Fill Parameter Estimates for Mean (μ)

### Sensitivity Analysis and Robustness of Findings

After conducting a sensitivity analysis, which involved removing outlier studies^41,42^ identified via the residual funnel plot (Figure 3a), the overall pooled effect remained robust. The sensitivity analysis yielded a pooled effect estimate of 0.505 (95% CI [0.401, 0.609]), corresponding to an estimated probability of a positive outcome of approximately 62% (95% PI [0.084, 0.926]). Heterogeneity among studies remained substantial, with a significant Q-test indicating variability across studies (Q (38) = 190.21, p < .001). The between-study variance was moderate, with τ² = 0.043 (τ = 0.208), while the I² statistic of 75.49% suggests that a large proportion of observed variability is due to real differences between studies rather than sampling error. The H² value of 4.08 further confirms considerable heterogeneity. Overall, the sensitivity analysis shows that removing high-effect outliers had minimal impact on the estimated effect, indicating that the original findings were robust. Despite some reduction in between-study variance, variability in intervention effects across studies remains substantial, as reflected in the wide prediction interval. This highlights the importance of contextual factors such as participant characteristics, intervention type, and implementation setting when interpreting the overall effectiveness of ASD interventions *(Table 6; Figure 7)*.

**Figure 7:**
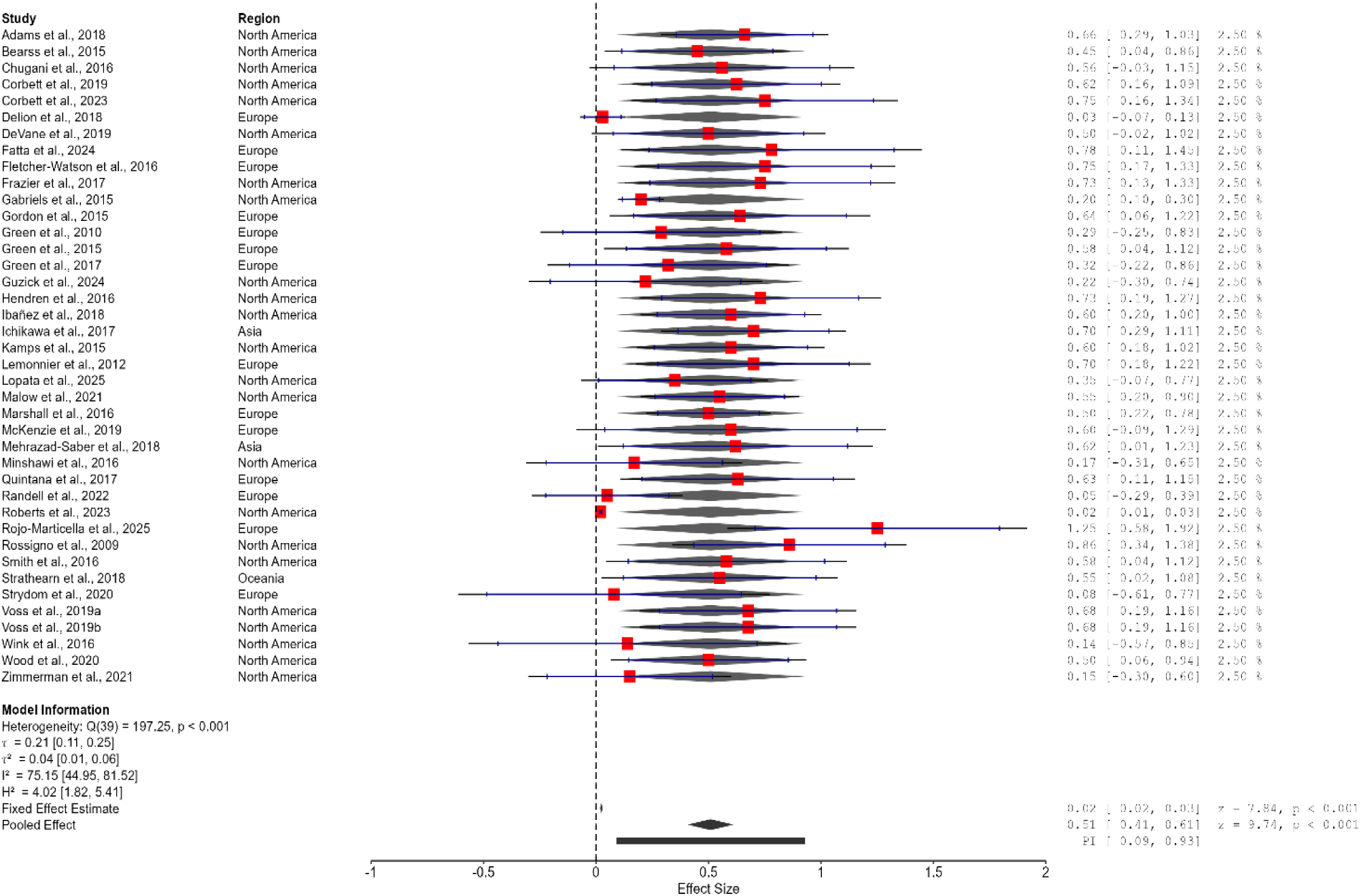
Sensitivity Analysis Forest plot of the pooled effect size and heterogeneity of interventions across studies (2004–2025).

**Table 6.**
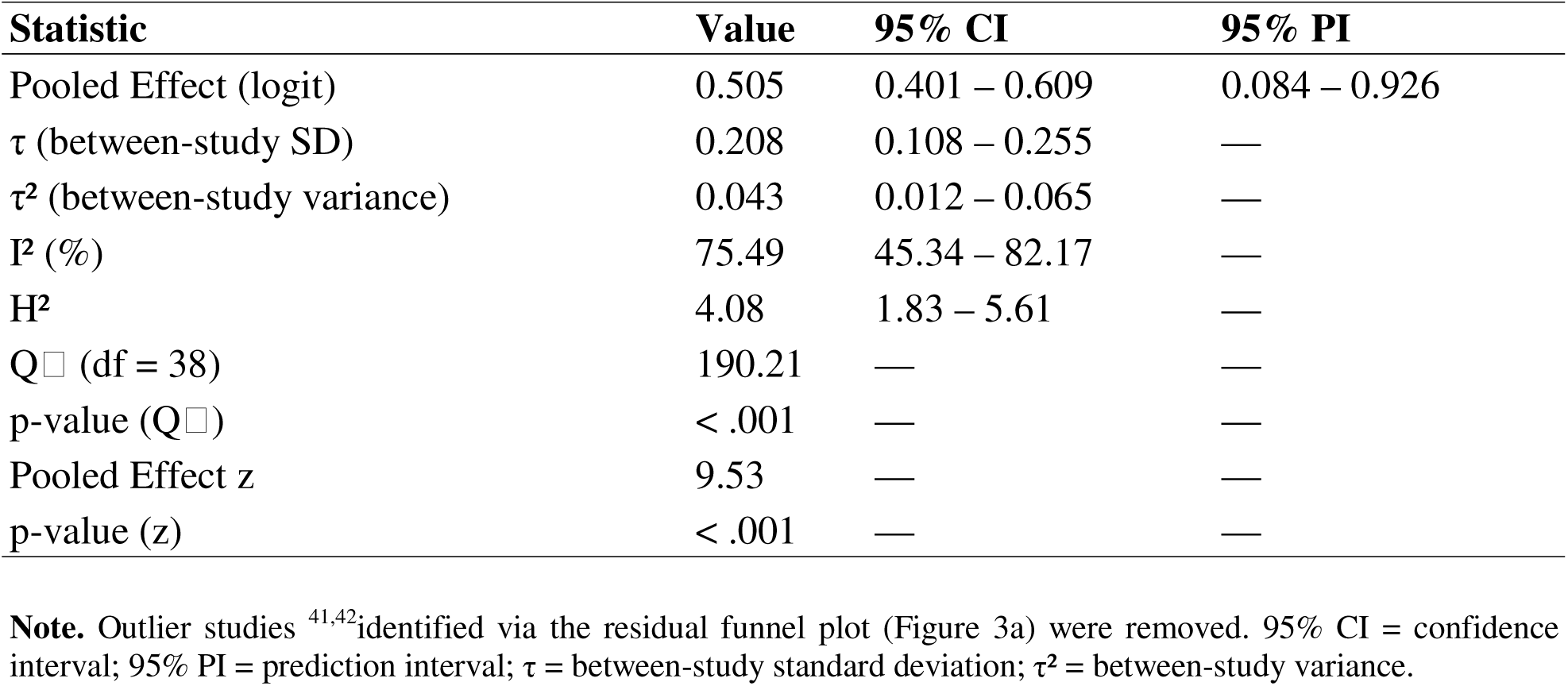
Sensitivity Analysis of Pooled ASD Intervention Effects (Outliers Removed)

| **Statistic** | **Value** | **95% CI** | **95% PI** |
| --- | --- | --- | --- |
| Pooled Effect (logit) | 0.505 | 0.401 – 0.609 | 0.084 – 0.926 |
| $\tau$ (between-study SD) | 0.208 | 0.108 – 0.255 | — |
| $\tau^2$ (between-study variance) | 0.043 | 0.012 – 0.065 | — |
| $I^2$ (%) | 75.49 | 45.34 – 82.17 | — |
| $H^2$ | 4.08 | 1.83 – 5.61 | — |
| $Q_{\chi^2}$ (df = 38) | 190.21 | — | — |
| p-value ($Q_{\chi^2}$) | < .001 | — | — |
| Pooled Effect z | 9.53 | — | — |
| p-value (z) | < .001 | — | — |

**Note.** Outlier studies <sup>41,42</sup> identified via the residual funnel plot (Figure 3a) were removed. 95% CI = confidence interval; 95% PI = prediction interval; $\tau$ = between-study standard deviation; $\tau^2$ = between-study variance.

### Frequentist Subgroup Analysis

#### a) By Intervention Category

The subgroup analysis showed variation in effectiveness across intervention categories. Nutritional and biomedical, behavioral and psychosocial, and pharmacological interventions all demonstrated moderate effects (64%, 63%, and 63%, respectively) (0.635, 95% CI [0.511, 0.744], PI [0.372, 0.837], p = .033, I² = 73.81%; 0.630, 95% CI [0.587, 0.671], PI [0.483, 0.756], p < .001, I² = 74.78%; 0.627, 95% CI [0.588, 0.665], p < .001, I² = 0%), although heterogeneity was high for nutritional and biomedical and behavioral and psychosocial interventions, while pharmacological interventions showed no heterogeneity. Digital and technology-based interventions had the highest pooled effect (67%) (0.672, 95% CI [0.598, 0.739], p < .001, I² = 0%), indicating a strong and highly consistent effect. In contrast, physical and occupational therapies showed a smaller, non-significant effect (52%) (0.523, 95% CI [0.484, 0.563], p = .249, I² = 63.35%), while combined pharmacological and behavioral interventions demonstrated a marginal effect (59%) (0.593, 95% CI [0.493, 0.685], p = .067, I² = 19.96%). Subgroup differences were statistically significant, Q[(5) = 22.63, p < .001, confirming variation across intervention types. Overall, although effect sizes were relatively similar across most categories, consistency varied considerably, with pharmacological and digital and technology-based interventions showing the most stable effects compared to the more heterogeneous nutritional and behavioral approaches *(Table 7; Figure 8)*.

**Figure 8:**
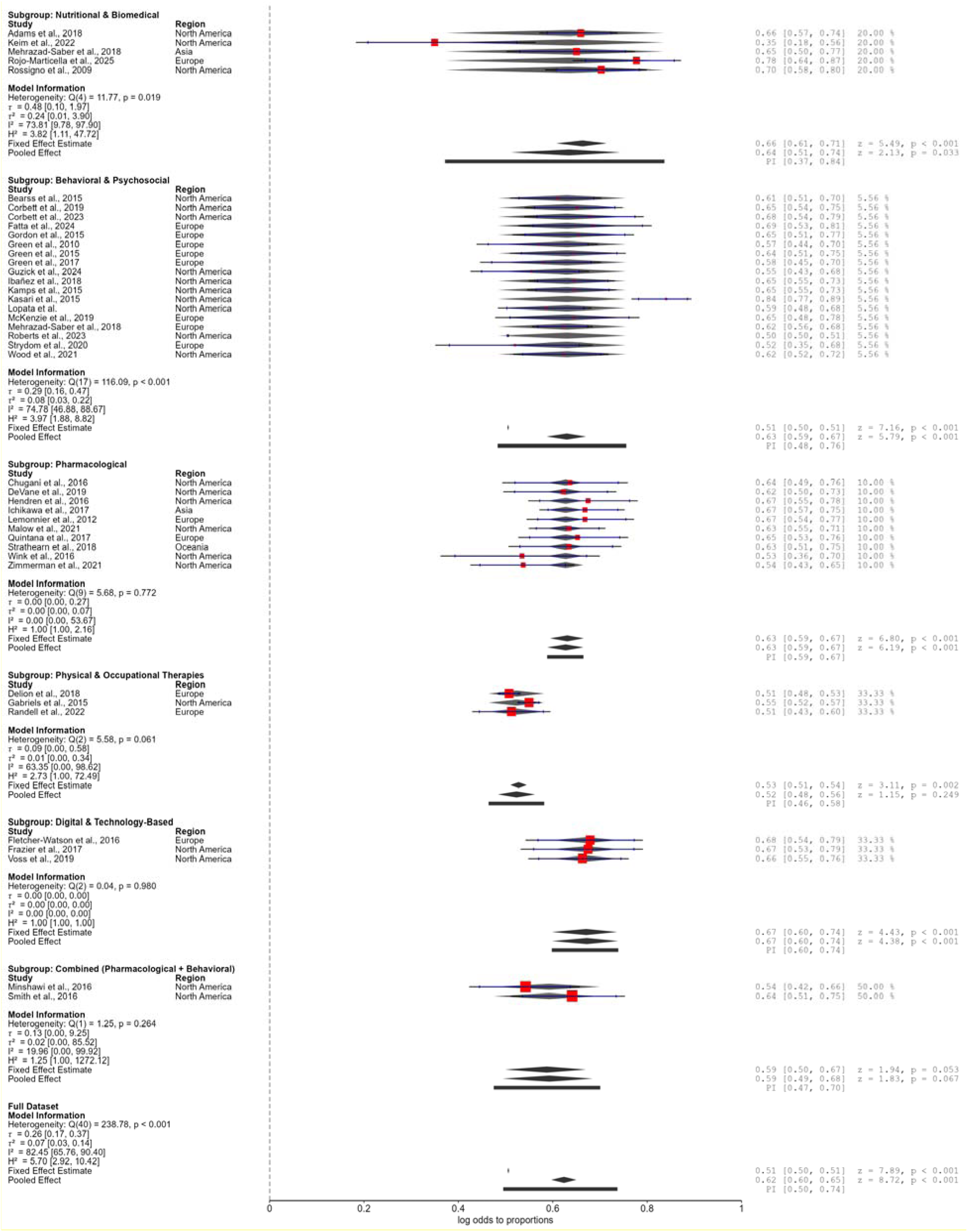
Forest plot and frequentist subgroup analysis of intervention effectiveness by intervention category (n = 41) **Note.** The figure presents forest plot–based meta-analytic estimates showing pooled effect sizes for each intervention category with corresponding 95% confidence intervals and test statistics. It also illustrates within-subgroup heterogeneity, highlighting variability in effect sizes across studies. In addition, the frequentist subgroup analysis displays pooled effect estimates (proportions) across intervention subgroups, demonstrating varying degrees of intervention effectiveness across categories.

**Figure 9.**
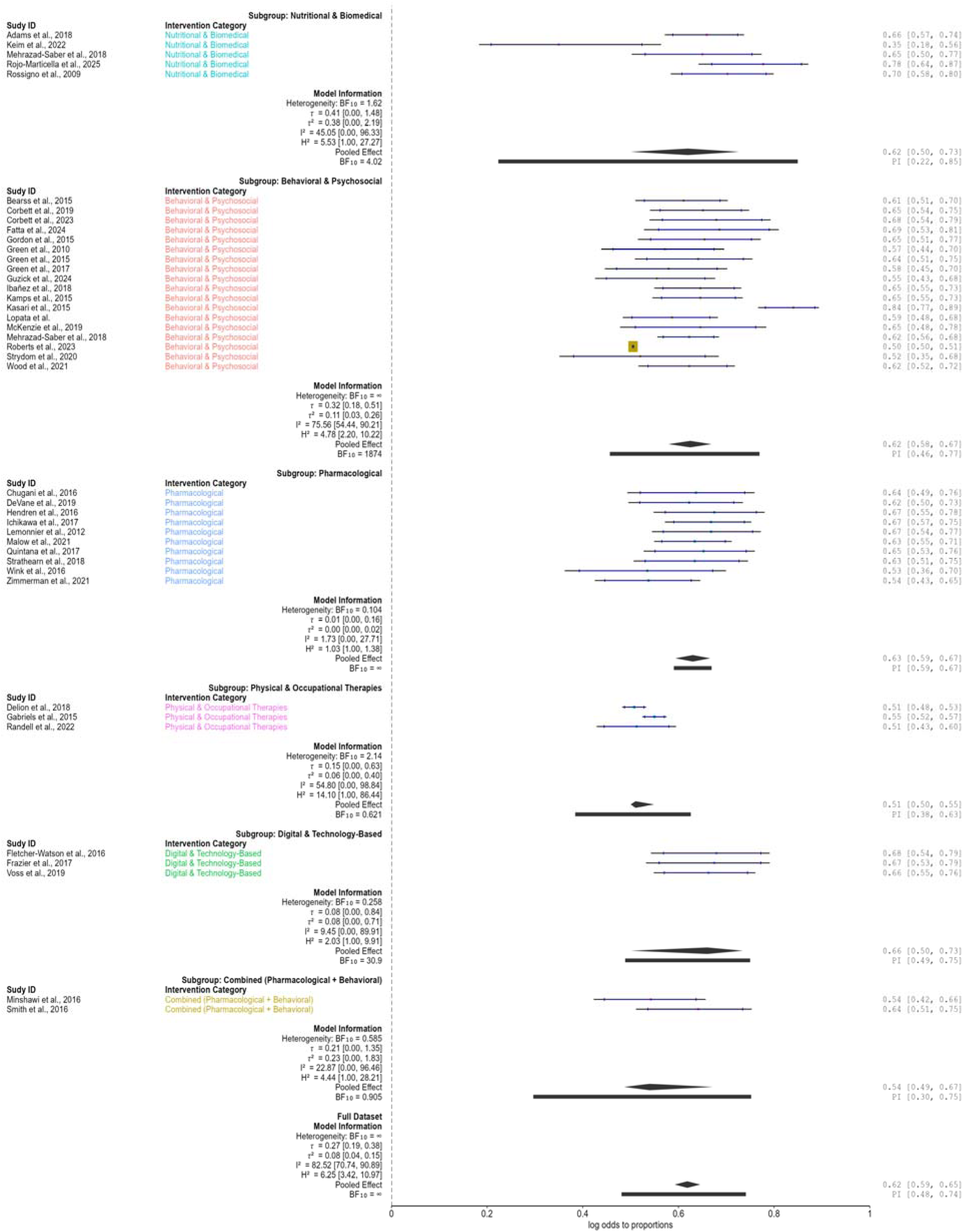
Forest plot of Bayesian subgroup meta-analysis by intervention type. **Note:** Squares represent pooled posterior mean effect estimates and horizontal lines indicate 95% credible intervals (CrIs); the vertical line denotes the null effect. Subgroups include behavioral/psychosocial, nutritional/biomedical, pharmacological, physical/occupational, digital/technology-based, and combined pharmacological–behavioral interventions. Bayes factors (BF) and heterogeneity (I²) are reported for each subgroup.

**Table 7.**
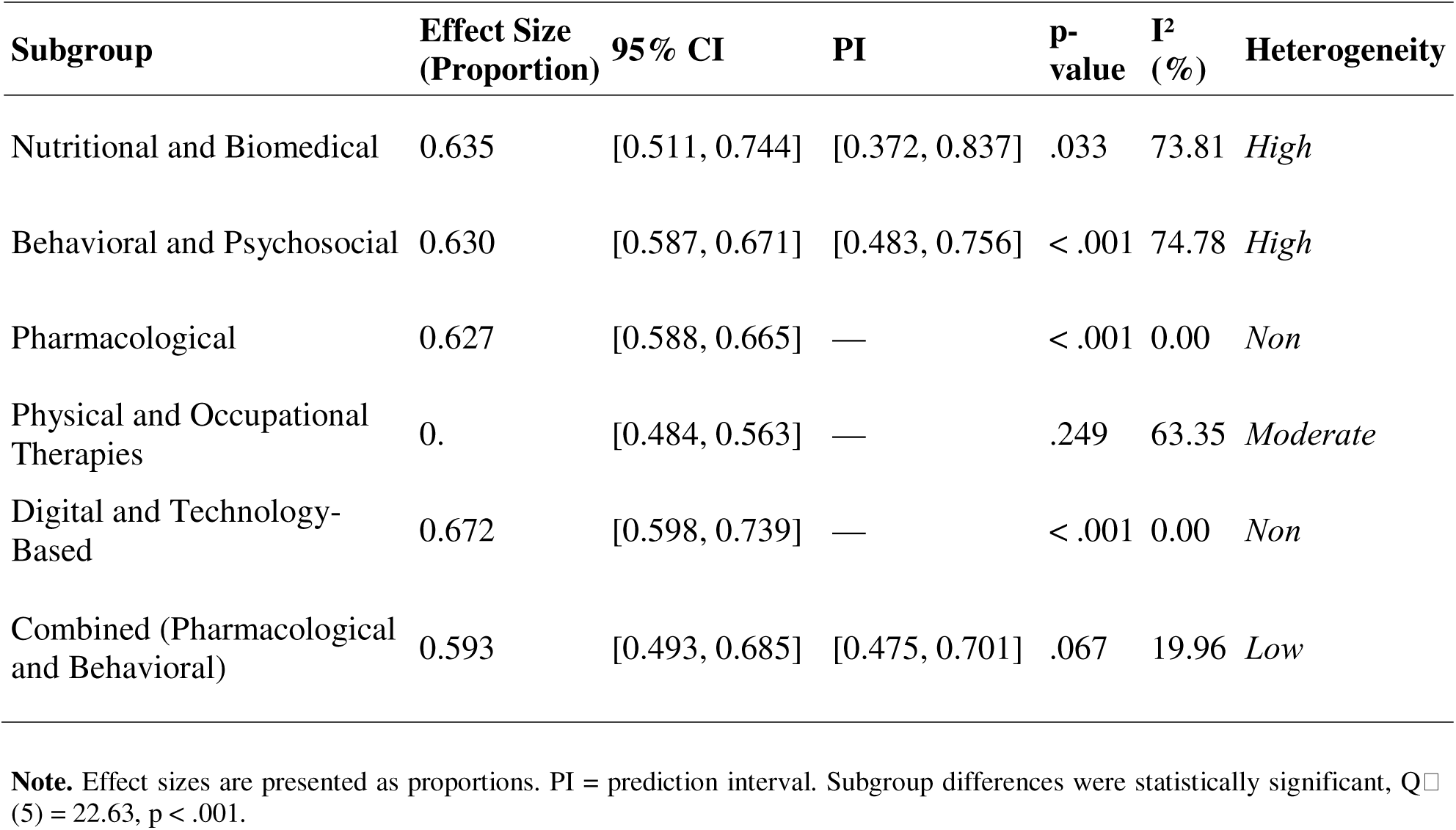
Subgroup Meta-Analytic Effect Estimates for Intervention Categories.

| **Subgroup** | **Effect Size**<br>**(Proportion)** | **95% CI** | **PI** | **p-**<br>**value** | **I<sup>2</sup>**<br>**(%)** | **Heterogeneity** |
| --- | --- | --- | --- | --- | --- | --- |
| Nutritional and Biomedical | 0.635 | [0.511, 0.744] | [0.372, 0.837] | .033 | 73.81 | *High* |
| Behavioral and Psychosocial | 0.630 | [0.587, 0.671] | [0.483, 0.756] | < .001 | 74.78 | *High* |
| Pharmacological | 0.627 | [0.588, 0.665] | — | < .001 | 0.00 | *Non* |
| Physical and Occupational<br>Therapies | 0. | [0.484, 0.563] | — | .249 | 63.35 | *Moderate* |
| Digital and Technology-Based | 0.672 | [0.598, 0.739] | — | < .001 | 0.00 | *Non* |
| Combined (Pharmacological<br>and Behavioral) | 0.593 | [0.493, 0.685] | [0.475, 0.701] | .067 | 19.96 | *Low* |

**Note.** Effect sizes are presented as proportions. PI = prediction interval. Subgroup differences were statistically significant, $Q(5) = 22.63$ , $p < .001$ .

#### b) By Geographic Region

The subgroup analysis examined variation in intervention effectiveness across geographic regions, including North America, Europe, and Asia. North America (62%) (0.619, 95% CI [0.582, 0.655], PI [0.475, 0.745], p < .001; I² = 84.21%) and Europe (63%) (0.626, 95% CI [0.579, 0.671], PI [0.495, 0.741], p < .001; I² = 62.34%) showed comparable moderate effects, although heterogeneity was substantial in North America and moderate in Europe. Asia (66%) (0.659, 95% CI [0.572, 0.737], p < .001; I² = 0%) demonstrated a similar pooled effect with no observed heterogeneity; however, this finding should be interpreted with caution due to the limited number of included studies. Subgroup differences were not statistically significant (Q (2) = 0.73, p = .694), indicating no evidence of variation in pooled effects across regions. The Oceania subgroup could not be estimated due to insufficient data. Overall, effect sizes were consistent in magnitude across regions, with variability largely driven by within-group heterogeneity, particularly in North America and Europe *(Table 8)*.

**Table 8.**
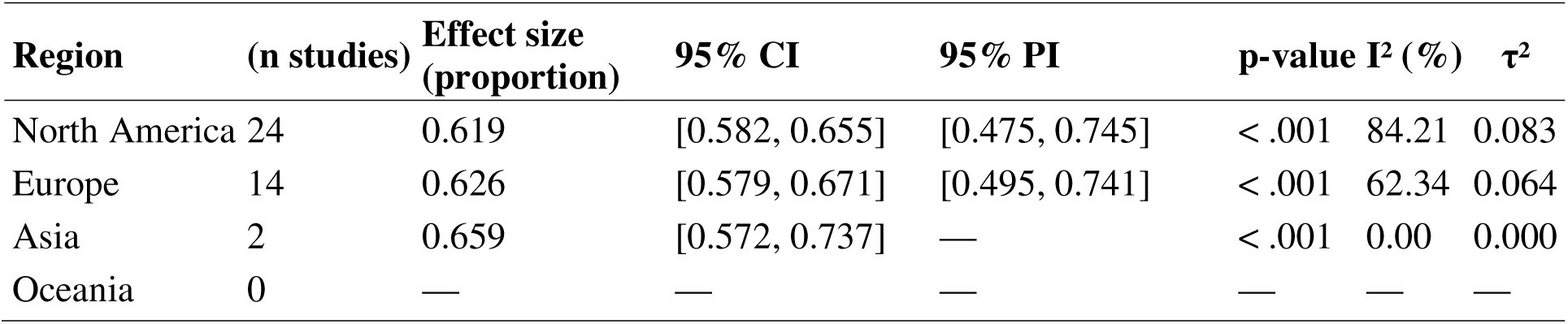
Frequentist Subgroup Meta-Analysis of Intervention Effectiveness by Geographic Region.

| Region | (n studies) | Effect size (proportion) | 95% CI | 95% PI | p-value | $I^2$ (%) | $\tau^2$ |
| --- | --- | --- | --- | --- | --- | --- | --- |
| North America | 24 | 0.619 | [0.582, 0.655] | [0.475, 0.745] | < .001 | 84.21 | 0.083 |
| Europe | 14 | 0.626 | [0.579, 0.671] | [0.495, 0.741] | < .001 | 62.34 | 0.064 |
| Asia | 2 | 0.659 | [0.572, 0.737] | — | < .001 | 0.00 | 0.000 |
| Oceania | 0 | — | — | — | — | — | — |

**Figure 9:** Frequentist Subgroup Meta-Analysis of Intervention Effectiveness by Geographic Region

**Note.** Effect sizes are presented as pooled proportions with 95% confidence intervals (CI) and prediction intervals (PI) where available. Oceania was excluded due to insufficient studies (fewer than two estimates). Subgroup differences were not statistically significant, Q[(2) = 0.73, p = .694.

#### c) By Age at Start of Intervention

The subgroup analysis examined variation by age at intervention onset, including Children, Youth, Adults, Children & Adolescents, Infants, and Toddlers. Children (49%) (0.486, 95% CI [0.364, 0.609], PI [0.098, 0.874], p < .001; I² = 48.06%) showed a moderate effect with moderate heterogeneity. Youth (41%) (0.406, 95% CI [0.119, 0.693], p = .006; I² = 0%) and Adults (49%) (0.487, 95% CI [0.136, 0.837], p = .006; I² ≈ 0%) yielded comparable effects with no observed heterogeneity, though both are based on few studies. Children and Adolescents (62%) (0.620, 95% CI [0.233, 1.007], PI [-0.178, 1.418], p = .002; I² = 76.66%) showed a larger but more variable effect, while Infants (45%) (0.450, 95% CI [0.067, 0.833], p = .021; I² = 0%) demonstrated a moderate effect with no heterogeneity. In contrast, Toddlers (84%) (0.840, 95% CI [-0.767, 2.447], PI [-1.924, 3.604], p = .306; I² = 97.86%) showed a highly variable and non-significant effect. Subgroup differences were not statistically significant (Q (5) = 0.98, p = .964), indicating no evidence of variation in pooled effects by age at intervention onset. Overall, effects were broadly comparable across age groups, though heterogeneity varied substantially, particularly among Children and Adolescents and Toddlers, suggesting important contextual and developmental influences *(Table 9)*.

**Table 9:** Frequentist Subgroup Meta-Analysis of Intervention Effectiveness by Age at Intervention Onset.

| Age group | n (studies) | Effect size (proportion) | 95% CI | 95% PI | p-value | I<sup>2</sup> (%) | τ<sup>2</sup> |
| --- | --- | --- | --- | --- | --- | --- | --- |
| a. Children | 25 | 0.617 | [0.587, 0.647] | [0.522, 0.705] | < .001 | 48.60 | 0.036 |
| b. Youth | 4 | 0.600 | [0.530, 0.667] | [0.530, 0.667] | .006 | 0.00 | 0.000 |
| c. Adults | 3 | 0.619 | [0.534, 0.698] | [0.534, 0.698] | .006 | 0.01 | 0.000 |
| d. Children & Adolescents | 5 | 0.650 | [0.558, 0.732] | [0.456, 0.805] | .002 | 76.66 | 0.127 |
| e. Infants | 2 | 0.611 | [0.517, 0.697] | [0.517, 0.697] | .021 | 0.00 | 0.000 |
| f. Toddlers | 2 | 0.698 | [0.317, 0.920] | [0.127, 0.974] | .306 | 97.86 | 1.316 |

**Note.** Effect sizes are pooled proportions with 95% confidence intervals (CI) and prediction intervals (PI) where available. Subgroup differences were not statistically significant, $Q(5) = 0.97$ , $p = .965$ . Estimates are based on log odds-to-proportion transformation.

### Bayesian Subgroup Meta-Analysis by Intervention Type

The Bayesian subgroup meta-analysis demonstrated consistent positive pooled effects across most intervention categories, although the strength of evidence and degree of heterogeneity varied substantially between subgroups. Behavioral and psychosocial interventions showed a moderate positive effect (62.5%) 0.625 (95% credible interval [CI]: 0.579–0.669) with decisive evidence for inclusion (BF ≈ 832.50), but heterogeneity was substantial (I² ≈ 75.6%), indicating considerable variability across studies. Nutritional and biomedical interventions also demonstrated a moderate positive effect (62.5%) 0.625 (95% CI: 0.500–0.728), with moderate evidence (BF = 4.67) and moderate heterogeneity (I² ≈ 40.3%). Pharmacological interventions produced a comparable moderate effect (63.0%) 0.630 (95% CI: 0.592–0.665) with decisive evidence (BF = ∞) and negligible heterogeneity (I² ≈ 1.7%), indicating highly consistent findings. Physical and occupational therapies showed a smaller effect (51.0%) 0.510 (95% CI: 0.498–0.551) with weak evidence against inclusion (BF = 0.63) and moderate to high heterogeneity (I² ≈ 55.9%). Digital and technology-based interventions demonstrated one of the strongest pooled effects (66.1%) 0.661 (95% CI: 0.500–0.736) with very strong evidence (BF ≈ 30.92) and low heterogeneity (I² ≈ 9.1%), indicating relatively consistent effects. Combined pharmacological and behavioral interventions yielded a moderate effect (53.9%) 0.539 (95% CI: 0.482–0.669), but with inconclusive evidence (BF = 0.92) and low to moderate heterogeneity (I² ≈ 23.5%), suggesting some uncertainty in their overall effectiveness *(Table 9)*.

### Meta-Regression Analyses

Meta-regression analyses indicated that intervention context (Q = 18.159, df = 8, p = .020), outcome domain (Q = 19.588, df = 6, p = .003), age at start (Q = 17.795, df = 5, p = .003), and intervention category (Q = 31.714, df = 5, p < .001) significantly moderated effect sizes, whereas follow-up duration (Q = 2.679, df = 5, p = .749) and intervention duration (Q = 1.777, df = 2, p = .411) did not. At the coefficient level, most contextual comparisons were associated with significantly larger effects relative to the reference category, including inpatient/clinical (B = 1.803, p = .002), community-based (B = 1.536, p = .003), school-based (B = 1.367, p = .003), hospital-based (B = 1.322, p = .041), home-based (B = 1.017, p = .002), and clinic-based settings (B = 1.033, p = .015). Outcome domain analyses showed no consistently significant individual contrasts, although adaptive functioning showed a trend toward larger effects (B = 0.659, p = .059). Age at start significantly moderated outcomes, with toddlers demonstrating larger effects (B = 1.326, p = .012), while other age groups were not significant. Intervention category also significantly moderated effects, with physical and occupational therapies (B = −1.230, p < .001) and combined pharmacological and behavioral interventions (B = −0.792, p = .009) associated with smaller effect sizes relative to the reference category. Overall, these findings suggest that heterogeneity in effect sizes is primarily explained by intervention context, age, and intervention type, whereas timing-related variables such as follow-up and duration did not significantly contribute to variability *(Table 10)*.

**Table 10.** Bayesian Subgroup Meta-Analysis by Intervention Type.

| Intervention type | Effect (Mean) | 95% CI | BF | I<sup>2</sup> (%) | Interpretation |
| --- | --- | --- | --- | --- | --- |
| Nutritional & Biomedical | 0.625 | 0.500–0.728 | 4.67 | 40.3 | *Moderate effect; moderate evidence; moderate heterogeneity* |
| Behavioral & Psychosocial | 0.625 | 0.579–0.669 | 832.50 | 75.6 | *Moderate effect; decisive evidence; substantial heterogeneity* |
| Pharmacological | 0.630 | 0.592–0.665 | ∞ | 1.7 | *Moderate effect; decisive evidence; negligible heterogeneity* |
| Physical & Occupational Therapies | 0.510 | 0.498–0.551 | 0.63 | 55.9 | *Small effect; weak evidence; moderate–high heterogeneity* |
| Digital & Technology-Based | 0.661 | 0.500–0.736 | 30.92 | 9.1 | ***Strongest effect; very strong evidence; low heterogeneity*** |
| Combined (Pharmacological + Behavioral) | 0.539 | 0.482–0.669 | 0.92 | 23.5 | *Moderate effect; inconclusive evidence; low–moderate heterogeneity* |

**Note.** CI = credible interval; BF = Bayes factor; I<sup>2</sup> = percentage of total variation due to heterogeneity. Values are derived from Bayesian meta-analytic models using log-odds transformed proportions.

**Table 11.** Meta-regression results examining moderators of intervention effect sizes.

| Predictor | $Q^2$ | df | p-value |
| --- | --- | --- | --- |
| Context | 18.159, $p = .020$ | 8 | .020 |
| Follow-up | 2.679, $p = .749$ | 5 | .749 |
| Outcome Domain | 19.588, $p = .003$ | 6 | .003 |
| Age at Start | 17.795, $p = .003$ | 5 | .003 |
| Intervention Duration | 1.777, $p = .411$ | 2 | .411 |
| Intervention Category | 31.714, $p < .001$ | 5 | < .001 |

**Note.** $Q^2$ = omnibus test statistic for moderator effects in meta-regression; df = degrees of freedom; p = p-value; fixed-effects meta-regression tests were conducted using a z-distribution.

## Discussion

### Principal Findings

The observed heterogeneity was likely attributable to multiple factors, including differences in intervention modalities, delivery settings, participant characteristics, outcome domains, and study design features. Diagnostic and analytical variability, as well as differences in implementation fidelity and follow-up duration, may have further contributed to the observed dispersion in effect sizes. Thus, while the overall evidence supports the effectiveness of ASD interventions, variability in outcomes—rather than the pooled estimate alone—emerges as a central finding of this synthesis, underscoring the context-dependent nature of intervention success, pin-pointing the need for careful consideration of population characteristics, intervention design, and delivery settings in both research and practice. This interpretation is further supported by the meta-regression analyses, which provide a formal test of whether study-level characteristics account for the observed heterogeneity. However, the meta-regression results indicated that heterogeneity was primarily explained by intervention context, age at intervention start, outcome domain, and intervention category, whereas follow-up duration and intervention duration were not statistically significant moderators, suggesting that not all study-level characteristics contributed equally to observed variability.

### Variation by Intervention Type and Population Context

Subgroup analyses showed broadly similar effectiveness across intervention types, with improvement rates of about 52% to 67%. Digital approaches performed best, while physical and occupational therapies were lowest. Although effect sizes were similar, variability remained high in several groups (I² = 63–75%). Pharmacological and digital interventions showed almost no variability, suggesting more consistent results. Regional outcomes were also similar (62–66%) with no major differences, though some variation still existed within regions. Age-based analyses followed the same pattern, but variability was higher, especially among toddlers. Overall, results were consistent across subgroups, but variation within them remained notable, pointing to the influence of context. These subgroup patterns are broadly consistent with the meta-regression findings, which identified intervention category as a significant source of heterogeneity, particularly highlighting reduced effects for physical and occupational therapies relative to other intervention types.

### Methodological and Contextual Moderators

Meta-regression identified key factors influencing outcomes, including setting, duration, outcome type, age, and intervention type. Omnibus tests indicated that context, outcome domain, age at start, and intervention category significantly explained heterogeneity (context: Q = 18.159, p = .020; outcome domain: Q□= 19.588, p = .003; age at start: Q□= 17.795, p = .003; intervention category: Q□= 31.714, p < .001), whereas follow-up duration and intervention duration were not significant moderators. Home-based and medium-duration (3–6 months) interventions showed larger effects, while clinic- and institution-based settings showed more variable and often smaller effects. Across contexts, most settings (e.g., inpatient/clinical, community-based, school-based) were associated with significantly higher effect sizes relative to the reference category, suggesting that intervention setting plays a key role in outcome magnitude. Family/psychosocial and adaptive outcomes, as well as interventions started in infancy, toddlerhood, and youth, showed lower effects. Age at intervention start was a significant moderator, with toddlers showing notably larger effects compared to the reference age group, while other age groups were not significant. Differences between intervention types also contributed to variations in results, with physical and occupational therapies and combined pharmacological–behavioral interventions associated with significantly smaller effect sizes, whereas other categories showed non-significant differences. However, interpretation of these coefficients should be made with caution, as variance inflation diagnostics indicated substantial multi-collinearity among predictors, suggesting that study-level characteristics were highly interrelated and limiting the ability to fully disentangle independent moderator effects. Consequently, observed associations may reflect overlapping contextual influences rather than isolated causal effects of individual moderators. Overall, effectiveness is strongly influenced by context and study design, with considerable unexplained variability still remaining.

### Sensitivity and Robustness

Sensitivity analyses showed that a few studies contributed heavily to variability, but removing them had little effect on the overall estimate (62%). Trim-and-fill analysis suggested some publication bias, lowering the estimate slightly to 59%, though results remained significant. Overall, the findings are robust, and the variability likely reflects real differences rather than just outliers. Importantly, the persistence of significant moderator effects in the meta-regression despite these adjustments suggests that observed heterogeneity is not solely attributable to outliers or publication bias, but reflects genuine variation across study contexts and intervention characteristics.

### Bayesian Interpretation

Bayesian analyses closely matched frequentist results (∼62–63%) and showed strong evidence of a positive effect. Although variability remained high, prediction intervals suggested that future studies are likely to show meaningful benefits. Convergence checks confirmed stable estimates. Overall, the Bayesian results support the effectiveness of ASD interventions while highlighting differences across contexts. Although Bayesian subgroup analyses were not formally modelled within the same multivariable framework, the consistency between Bayesian and frequentist subgroup patterns provides converging evidence that intervention category and contextual factors are key sources of variation in intervention effects.

### Comparison with Prior Literature

Prior meta-analyses of interventions for Autism Spectrum Disorder (ASD) have consistently reported moderate positive effects across a range of therapeutic approaches, broadly aligning with the present pooled estimate indicating that approximately 62% of participants benefit from intervention. For example, earlier syntheses of behavioral and developmental interventions have reported effect sizes in the small-to-moderate range, particularly for early intensive behavioral interventions and parent-mediated approaches^70–73^. Similarly, umbrella reviews and large-scale evidence syntheses have concluded that, while many ASD interventions demonstrate efficacy, the magnitude of benefit is often variable and context-dependent^74,75^. The present findings extend this literature by integrating both frequentist and Bayesian approaches across a larger and more methodologically diverse evidence base, providing convergent evidence of a moderate-to-large overall effect.

Consistent with prior research, the current analysis identified substantial between-study heterogeneity (I² ≈ 75–82%), reinforcing longstanding concerns regarding variability in intervention outcomes across studies ^9,76^. Previous meta-analyses have attributed such heterogeneity to differences in intervention intensity, fidelity, participant characteristics, and outcome measurement, particularly in complex, multi-component interventions^72,77^. The wide prediction intervals observed in the present study (∼50–73%) further support the interpretation that intervention effectiveness is not uniform, but varies meaningfully across contexts. This aligns with earlier work emphasizing that pooled estimates in ASD intervention research should be interpreted as average effects across heterogeneous conditions rather than universally applicable benchmarks^78,79^.

Subgroup findings in the current study are also broadly consistent with prior literature. Behavioral and psychosocial interventions demonstrated moderate effectiveness, in line with established evidence supporting applied behavior analysis and related approaches^71,80^. Pharmacological interventions showed comparable average effects but lower heterogeneity, reflecting findings from systematic reviews indicating more standardized protocols and outcome measures in medication trials^81–84^. Notably, digital and technology-based interventions demonstrated relatively strong and consistent effects, supporting emerging evidence that technology-assisted therapies may enhance engagement and scalability, although this remains an evolving area of research^85,86^. In contrast, physical and occupational therapies showed smaller and more variable effects, consistent with previous reviews highlighting mixed evidence and methodological limitations in these domains^87,88^.

The identification of contextual and methodological moderators in the present analysis further aligns with prior findings. Previous studies have shown that intervention setting, duration, and delivery format significantly influence outcomes, with home-based and parent-mediated interventions often yielding stronger or more generalizable effects^41,89,90^ [3,13]. The present results similarly indicate that home-based and medium-duration interventions are associated with greater effectiveness, while clinic- and institution-based programs tend to yield smaller effects. Moreover, variability across outcome domains observed here reflects ongoing challenges in measuring complex constructs such as adaptive functioning and psychosocial outcomes, which have been noted as less responsive or more difficult to standardize in prior research^73,91,92^.

The robustness of the present findings, demonstrated through sensitivity analyses and trim-and-fill adjustments, is also consistent with earlier meta-analyses showing that while publication bias and small-study effects may modestly inflate effect sizes, they rarely negate the overall conclusion of intervention benefit^93–95^. The close agreement between frequentist and Bayesian estimates in the current study further strengthens confidence in the results, echoing recent methodological work advocating for Bayesian approaches to better characterize uncertainty and heterogeneity in ASD research^96–98^.

Overall, the findings of this meta-analysis are consistent with previous research. The results indicate moderate effectiveness, with considerable variation across studies. This variation appears to be influenced by contextual, demographic, and intervention-specific factors, as confirmed by the meta-regression findings. Together, these results suggest that outcomes of ASD interventions are not determined by a single superior approach, but rather by how, where, and for whom the intervention is implemented. Therefore, context-sensitive and individualized intervention strategies are essential. However, given evidence of substantial multi-collinearity among moderator variables, these findings should be interpreted as reflecting overlapping and interdependent contextual influences rather than fully independent effects of individual study-level characteristics.

## Data Availability

All data produced in the present study are available upon reasonable request to the authors

